# Effects of speculum lubrication on cervical smears for cervical cancer screening: a double blind randomized clinical trial

**DOI:** 10.1101/2023.09.17.23295694

**Authors:** Chito P. Ilika, George U. Eleje, Michael E. Chiemeka, Frances N. Ilika, Joseph I. Ikechebelu, Valentine C. Ilika, Emmanuel O. Ugwu, Ifeanyichukwu J. Ofor, Onyecherelam M. Ogelle, Osita S. Umeononihu, Johnbosco E. Mamah, Chinedu L. Olisa, Chijioke O. Ezeigwe, Malarchy E. Nwankwo, Chukwuemeka J. Ofojebe, Chidinma C. Okafor, Onyeka C. Ekwebene, Obinna K. Nnabuchi, Chigozie G. Okafor

## Abstract

**Background:** Speculum lubrication may help to reduce the pain experienced during Pap-smears collection and hence increase uptake of cervical cancer screening and repeat testing but there are fears of its interference with cytological results.

**Aim:** To determine and compare adequacy of cervical cytology smears and mean pain scores of women undergoing cervical cancer screening with or without speculum lubrication.

**Methods:** This was a randomized controlled study of 132 women having cervical cancer screening at a tertiary hospital in Nigeria. Sixty-six participants each were randomly assigned to the ‘Gel’ and ‘No Gel’ groups respectively. Pap-smears were collected from each participant with lubricated speculum (‘Gel group’) or non-lubricated speculum (‘No Gel group’). The primary outcome measures were; the proportion of women with unsatisfactory cervical cytology smears and the mean numeric rating scale pain scores while the secondary outcome measures were the proportion of women that are willing to come for repeat testing and the cytological diagnosis of Pap-smear results.

**Results:** The baseline socio-demographic variables were similar in both groups. There was no significant difference in the proportion of unsatisfactory cervical smear results between the two groups (13.6% vs. 21.2% p = 0.359). However, the mean pain scores were significantly lower in the gel group than in the no gel group (45.04 vs 87.96; p<0.001). An equal proportion of the participants in each group (90.9% vs. 90.9%; p>0.999) were willing to come for repeat cervical smears in the future.

**Conclusion:** Speculum lubrication did not affect the adequacy of cervical-smears but significantly reduced the pain experienced during Pap-smear collection. Also, it did not significantly affect willingness to come for repeat cervical smears in the future.

The Trial was registered with **Pan-African Clinical Trial Registry** with unique identification/registration no: **PACTR2020077533364675.**

## Introduction

Cancer of the cervix is the fourth most prevalent malignancies in females with a projected incidence of 570,000 in 2018.^1^ It represents 6.6 per cent of all gynaecological carcinomas and is responsible for 7.5 per cent of all female malignancy mortality worlwide.^1^ Out of the 311 000 cases of mortality estimated from carcinoma of the cervix yearly, greater than 85% of these deaths happen in areas that are less developed^1^.

Introducing screening for cancer of the cervix and improving its uptake especially in developing countries is key to decreasing the death burdens from malignancy of the cervix. In addition, the usage of screening programs can be improved via increasing sensitization on the risk factors of carcinoma of the cervix which include early coitarche, multiple sexual partners, HPV infections, extensive use of oral contraceptives and HIV infections^2^.

The conventional Papanicolaou test remains the commonest screening method for cancer of the cervix^3^. In high income countries, there are well structured programs which enable vaccination of girls against HPV and regular screening of women resulting in remarkable decrease in the development and complications of carcinoma of the cervix^4,5,6^. Screening helps to achieve early identification and intervention which prevents up to 80% of cervical malignancies in high income countries,^4,5,6^. whereas in poor income countries, there is poor access to vaccination and testing resulting in diagnosis of cervical cancer mainly in advanced stages^7,8^. In addition, these countries lack access to facilities required for the management of such advanced stage diseases giving rise to a greater death rate from malignancy of the cervix in poor countries^7,8^.

In as much as efforts are being made to improve vaccinations for HPV in order to prevent carcinoma of the cervix, early identification of premalignant lesions of the cervix through cervical Papanicolaou smear cytology screening remains a key factor for achieving a decline in the development and complications of carcinoma of the cervix in low and middle income countries where vaccination for Human Papilloma Virus services is limited^9^.

The Papanicolaou test is a well-recognized, efficient and reliable tool employed in early identification of premalignant lesions of the cervix resulting in substantial decrease in disease burden of carcinoma of the cervix^10^. It is cost-effective and the technique is simple. Despite this, the uptake of Papanicolaou smear in our environment remains poor, Pain and discomfort associated with examination of the vagina can discourage women from assessing regular test, other factors that may hinder compliance include lack of awareness, cost implication, anxiety and cultural beliefs^7,11^.

Insertion of speculum for examination of vagina is an important factor responsible for non-compliance to regular screening and repeat testing for carcinoma of the cervix because of the embarrassment, anxiety, pain and discomfort associated with it^12^. In Australia, a research seeking to find out the attitude of women concerning self-insertion compared to physician insertion of the speculum showed that 91% of the study population will prefer to insert the speculum by themselves rather than a physician doing it because of the embarrassment and discomfort associated with it^13^.

During intercourse, lubrication is physiologically essential for easy penetration of the vagina and absence of optimal lubrication results in dyspareunia. So we cannot justify inserting without lubrication a rigid instrument like a speculum into the vagina. Speculum lubrication should be employed to minimize discomfort and pain during vaginal examinations thereby ultimately increasing compliance for screening. However, applying lubricating gel on the vagina is not encouraged by gynaecologic literature, also students and resident doctors in training are advised against lubricating the speculum while collecting sample due to the worry that it may interfere with cytology results of cervical smears often leading to inadequacy.^10,14^ However, there is paucity of convincing evidence to prove that using lubricating gel can prevent proper cytological analysis^15,16,17^.

The aim of this study was to determine the effects of speculum lubrication on the adequacy of cervical cytology smears and to determine if it decreases pain/discomfort in women undergoing cervical cancer screening by means of cervical smears and also compare the proportion of women willing to come for repeat cervical smears in the future.

## Materials and Methods

This work was a prospective randomized controlled trial of 132 women having cervical cancer screening at the gynaecologic clinic of Nnamdi Azikiwe University Teaching Hospital (NAUTH) Nnewi, Anambra State, Nigeria from 21^st^ Augusts, 2020 to 31^st^ December, 2020. Ethical approval was obtained from NAUTH Ethics review committee with the approval reference number: NAUTH /CS/66/VOL.12/098/2019/040.

The participants included pre-menopausal women of atleast 25years and post-menopausal women who required pap smear screening. Women excluded were virgins, recent sexual intercourse, pregnant women, women having their menstrual period, women with any overt cervical pathology and/or current treatment of any vaginal condition, women with vulvar pathologies or those on hormone replacement therapy. Women with vaginitis, those undergoing vulvectomy or vaginectomy and women who had fertility-sparing surgery were also excluded.

The sample size was calculated using the formula for sample size determination of minimum sample size for experimental studies (Difference in proportions). Substituting for the values of proportion of unsatisfactory smears as found in a previous study ^11^

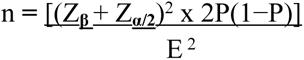

Where P **=** (P_1_ + P_2_) / 2 **=** (0.01+ 0.005) / 2 **=** 0.0075, E **=** Effect size **=** P_1_ - P_2_ **=** 0.01-0.005 **=** 0.005, Z**_β_ =** Corresponding Z value at 80% Power = 0.84. Z**_α/2_ =** 1.96 (Corresponding Z value at 95% confidence level) Adjusting for finite population, N = 60 samples from clinic in 3 months.

n_s_ **=** n^1^ / 1 + n^1^/N, where; n_s_ **=** Adjusted sample size, n^1^ **=** Calculated sample size, N **=** population for study. Considering attrition rate of 10.0%, the total number of subjects was 132 (66 in each arm).

The procedure was explained and participants counselled on the procedure, pre-procedure precautions like abstinence from sexual intercourse for atleast 24 hours prior to the procedure and avoidance of douching were observed. Trained research assistants (Senior Registrars) and the researchers were involved in counselling of participants and collection of the pap smears. Following a written informed consent, selected women were randomly assigned to the gel group or no gel group. The sequence of randomization was generated by the computer via randomly permuted blocks (blocks of 4, allocation ratio 1:1). To keep a similar number of subjects in each group, a block randomization method with blocks of 4 was used. An independent person who was not involved in the study performed the randomization using computer software program available at http://mahmoodsaghaei.tripod.com/Softwares/randalloc.html. Allocation concealment was done using serially numbered opaque sealed envelopes which contained either a sheet of paper showing gel group (Group A) placing the participant into the group using the speculum lubrication or a paper displaying no gel group (group B) placing the participant into the group receiving dry speculum (without speculum lubrication). The envelopes were kept and opened by an independent person. Once the envelope was opened the allocation of the participant was not changed. The histopathologist and participants were blinded and so were not aware of the intervention arm hence double blind.

The study procedure required the participants to be placed in dorsal position with the legs flexed at the knee and adducted at the hips. Following identification of appropriate size speculum, 2mls of KY jelly (Dionel, Maryland -USA) was applied on the entire external surfaces of the metal Cusco’s speculum and inserted gently into the vagina to visualize the cervix. A non-lubricated speculum was used for the control group. A dry gauze swab was used to clear any mucous on the cervix. A cytobrush (Liqui-PREP^TM^ LGM International, Inc. Melbourne USA) was used to take the sample for Pap smear. Having obtained the sample, the head of the brush was removed and put into the liquid preparative collection vial containing a liquid based medium. (Liqui-PREP^TM^ LGM International, Inc. Melbourne USA) The specimen in the vial was then carefully mixed in the liquid based medium. Then, the speculum was gently withdrawn. The patient was cleaned and counselled on the outcome. After each smear the patient was asked to rate her pain at the end of the procedure using a Numeric Rating Scale for pain from 0 (no discomfort) to 10 (most discomfort) indicate if she was willing to come for repeat testing in the future. Numeric Rating Scale for pain was used because it is authenticity and its universal acceptance for evaluation of pain.^11^

At histopathology laboratory of NAUTH, the vial was transferred into a centrifuge tube with same quantity of cleaning solution. The fluid was then centrifuged for 10 minutes at 1400 rpm; the supernatant was removed. Smears were made from the sediments after mixing with the cellular base solution (Liqui-PREP^TM^ LGM International, Inc. Melbourne USA), stained by the Papanicolaou staining method and reported using the Bethesda system stating whether the sample was satisfactory or not and the reason for unsatisfactory results. Samples were recorded as “unsatisfactory” if 75% of the cells were obscured by blood or inflammation, or if drying artifact or gel overlay were present. Otherwise, they were recorded as “satisfactory”.^10,18^. The patients with unsatisfactory smear results and positive cytological result were referred to the managing gynaecological unit for management and follow up while those with negative cytological result were counselled for repeat testing.

The primary outcome measures were; The proportion of women with unsatisfactory cervical cytology smears and the mean numeric rating scale pain scores. While the secondary outcome measures were; The proportion of women that are willing to come for repeat testing and the cytological diagnosis of Pap smear results.

Statistical Packages for Social Sciences (SPSS) IBM Corp version 25.0 was employed in the analysis of results. Tables were used to represent collected data. Means and standard deviations were used to represent continuous data. Mann-Whitney U Test was used to assess non-parametrics variables and chi squared test was used for categorical variables. Statistical significance was deduced at p-value less than 0.05.

## Results

149 women were assessed for eligibility, however, 132 participants met the inclusion criteria and were randomized into the Gel group (n=66) and No gel group (n=66). A flow diagram describing the participants flow through the study is shown in Figure 1. Table 1 shows the socio-demographic characteristics of participants in both groups. In terms of educational qualification, 89 (67.4%) of the participants had tertiary education while 28% and 4.5% attained secondary and primary education respectively. There was no significant difference in the baseline sociodemographic data of the two groups.

**Figure 1:**
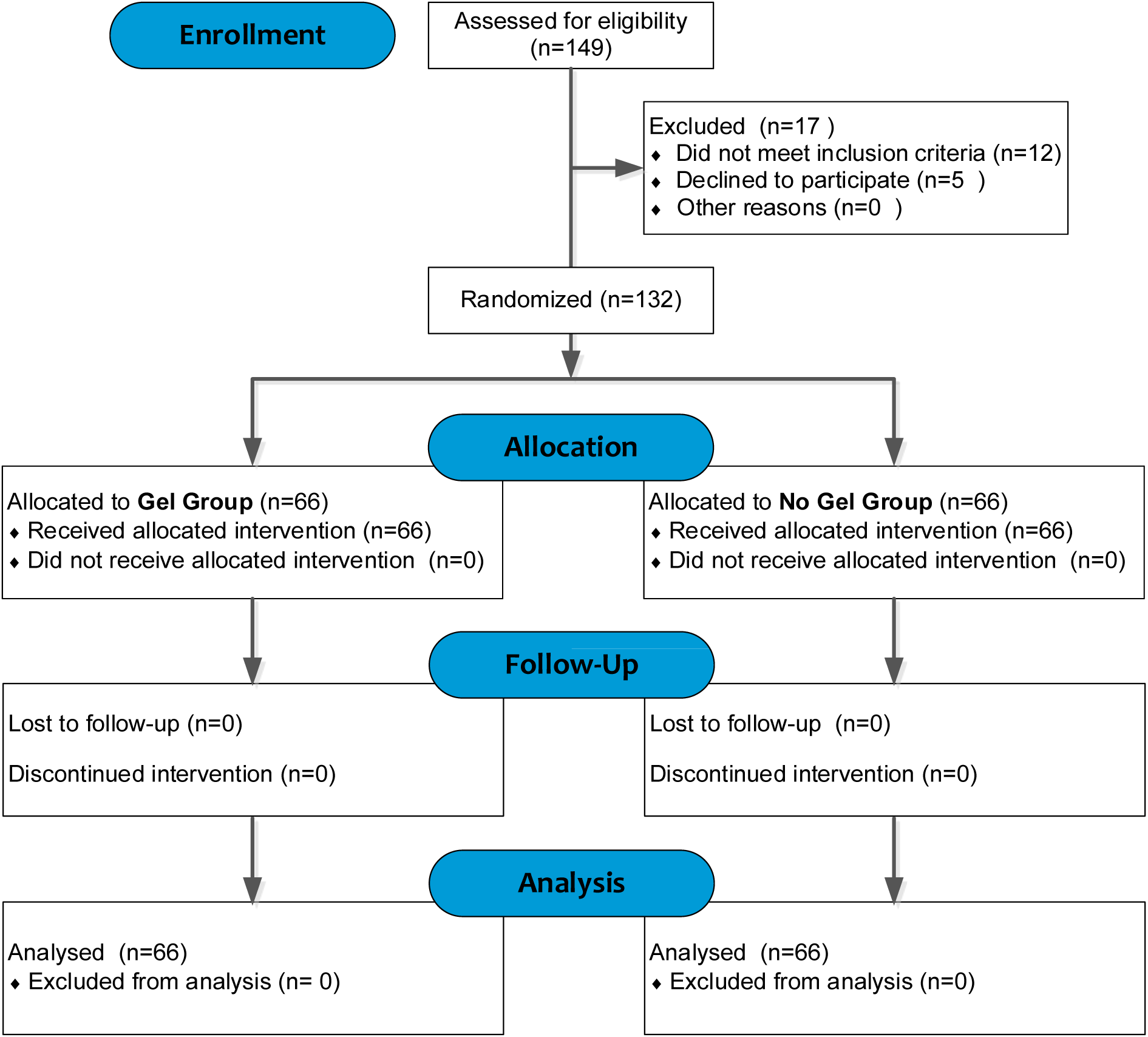
Consort flowchart for study participants

**Table 1:**
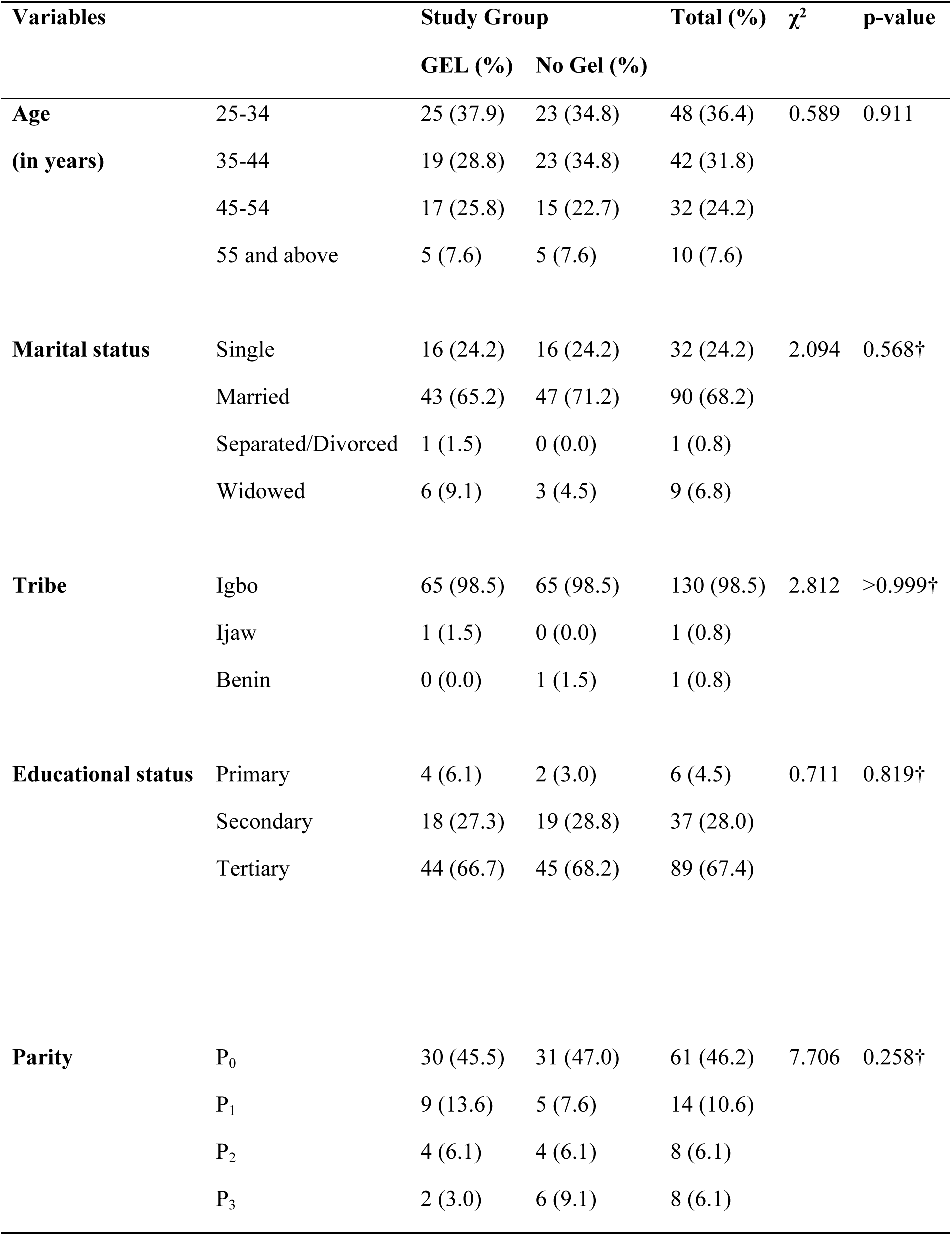
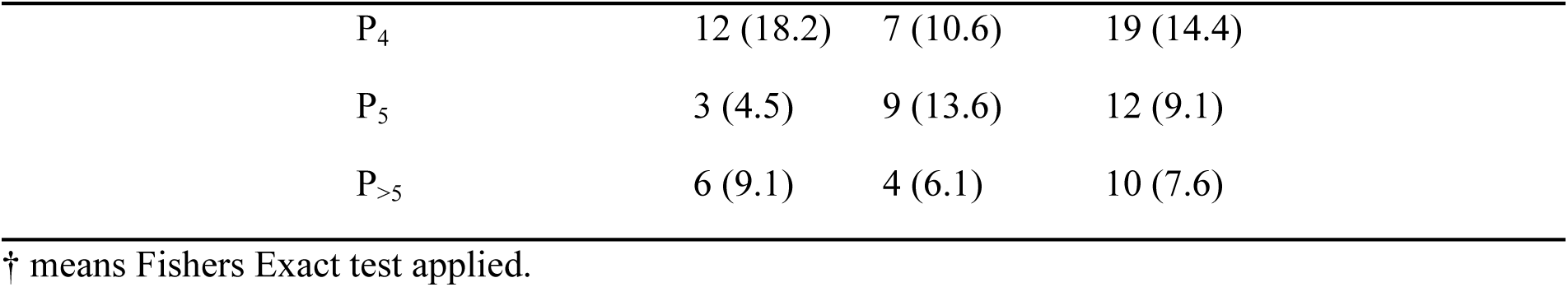
Socio-demographic characteristics of the participants.

There was no significant difference in the proportion of unsatisfactory cytology smear (13.6% vs 21.2%; p=0.359) and normal Pap smear results (100.0% vs 98.1%; p=0.477) in the gel group and no gel group, respectively. This is shown in Table 2.

**Table 2:** Comparison of cytology smear adequacy and cytological diagnosis of Pap Smears results among the study participants.

| Variables | | Study Group | | Total (%) | $\chi^2$ | p-value |
| --- | --- | --- | --- | --- | --- | --- |
|  |  | GEL (%) | No Gel (%) |  |  |  |
| **Smear sample status** | Satisfactory | 57 (86.4) | 52 (78.8) | 109 (82.6) | 1.316 | 0.359 |
|  | Unsatisfactory | 9 (13.6) | 14 (21.2) | 23 (17.4) |  |  |
| **PAP smear result** | LSIL | 0 (0.0) | 1 (1.9) | 1 (0.9) | 1.106 | 0.477† |
|  | NILM | 57 (100.0) | 51 (98.1) | 108 (99.1) |  |  |

† Fisher's Exact test used. LSIL = Low Grade Squamous Intraepithelial Lesion; NILM= Negative for Intraepithelial lesion or malignancy

The reasons for the unsatisfactory smear results were due to obscurity by marked inflammation (11.1% vs 14.3%; p=0.691) and presence of scanty cells (88.9% vs 85.7%; p=0.691) in the Gel and No Gel group respectively. Drying artifact due to gel was not reported by the histopathologist. This is shown in Table 3.

**Table 3:**
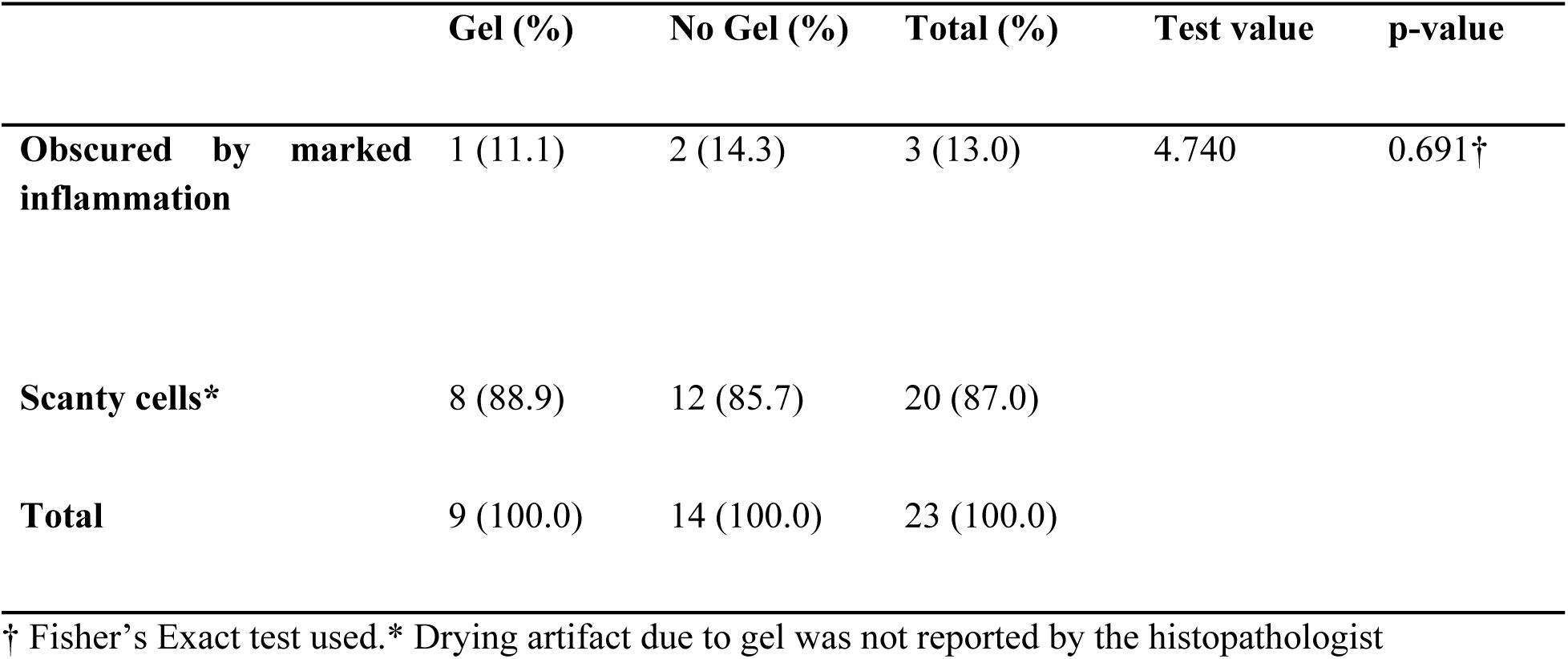
Comparison of the reasons for unsatisfactory smear results as reported by the histopathologist.

The mean pain scores were statistically lower in the Gel group than in the No gel group (45.04 vs 87.96; p<0.001). While 3 people from the no gel group had scores of 9 and 10, the pain scores in the gel group were less than 9. Additionally, 69.7%, 22.7%, and 7.6% of the women in the gel group had mild, moderate and severe pain respectively, while 3.0%, 68.2%, and 18.2% in the no gel group had mild moderate and severe pain respectively (p<0.001). This is shown in Table 4 and 5.

**Table 4:**
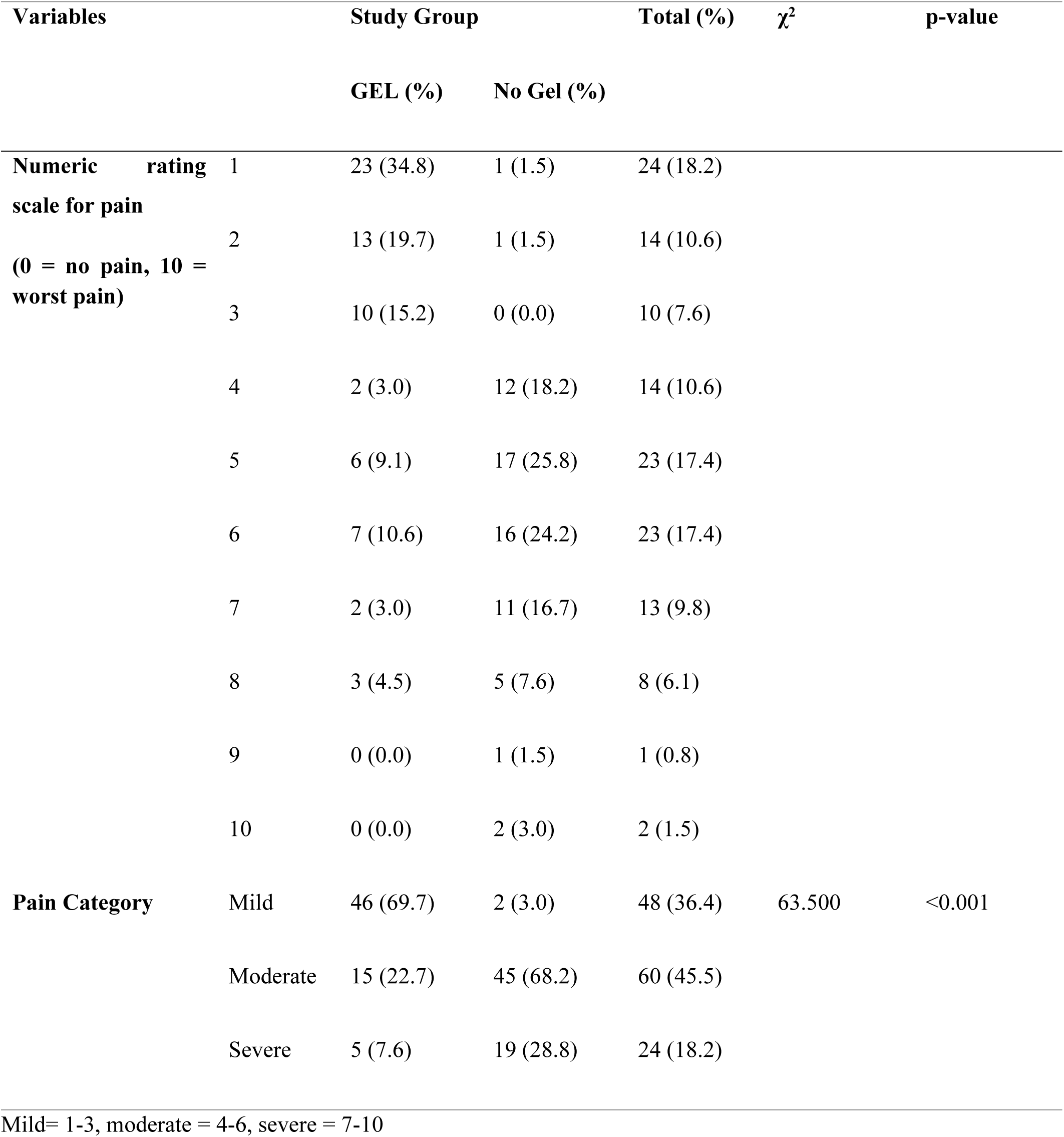
Comparison of Pain scores of the participants.

**Table 5:**
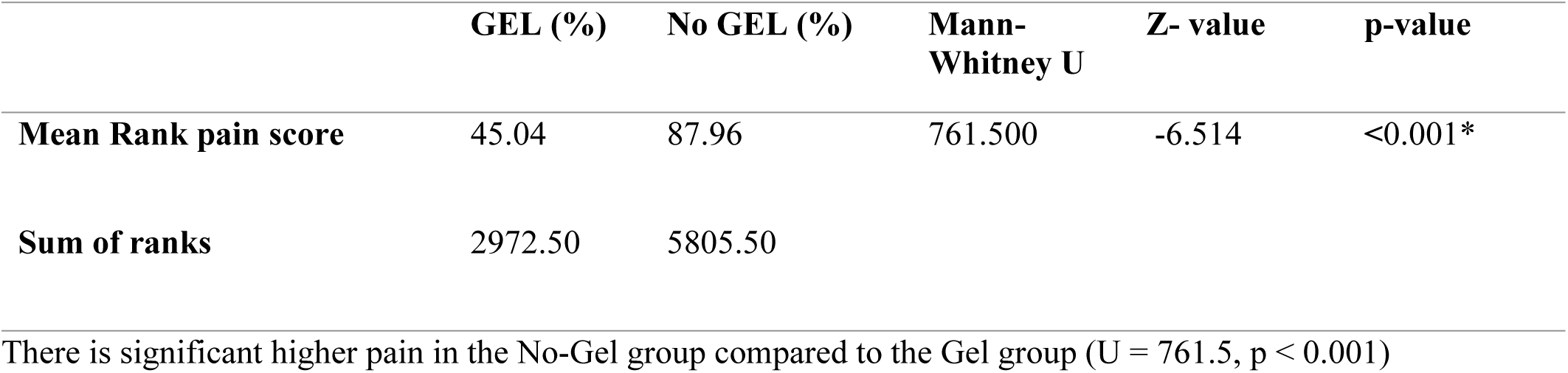
Comparison of Pain scores of the participants using Mann-Whitney U Test.

|  | GEL (%) | No GEL (%) | Mann-Whitney U | Z- value | p-value |
| --- | --- | --- | --- | --- | --- |
| Mean Rank pain score | 45.04 | 87.96 | 761.500 | -6.514 | <0.001* |
| Sum of ranks | 2972.50 | 5805.50 |  |  |  |

There is significant higher pain in the No-Gel group compared to the Gel group (U = 761.5, p < 0.001)

The association between category of pain and parity among the participants is shown in Table 6. The pain scores appear to be more at the extremes of parity.

**Table 6:** Association between category of pain and parity among the participants.

| Parity | Pain Category | Study Arms (%) | | $\chi^2$ | p-value |
| --- | --- | --- | --- | --- | --- |
|  |  | Gel | No Gel |  |  |
| **P<sub>0</sub>** | Mild | 21 (70.0) | 2 (6.5) | 26.216 | **<0.001** |
|  | Moderate | 6 (20.0) | 19 (61.3) |  |  |
|  | Severe | 3 (10.0) | 10 (32.3) |  |  |
| **P<sub>1</sub>** | Mild | 6 (66.7) | 0 (0.0) | 6.031† | 0.056 |
|  | Moderate | 2 (22.2) | 4 (80.0) |  |  |
|  | Severe | 1 (11.1) | 1 (20.0) |  |  |
| **P<sub>2</sub>** | Mild | 2 (50.0) | 0 (0.0) | 2.766† | 0.429 |
|  | Moderate | 2 (50.0) | 3 (75.0) |  |  |
|  | Severe | 0 (0.0) | 1 ((25.0) |  |  |
| **P<sub>3</sub>** | Mild | 1 (50.0) | 0 (0.0) | 2.895† | 0.286 |
|  | Moderate | 1 (50.0) | 4 (66.7) |  |  |
|  | Severe | 0 (0.0) | 2 (33.3) |  |  |
| **P<sub>4</sub>** | Mild | 9 (75.0) | 0 (0.0) | 10.276† | **0.003** |
|  | Moderate | 2 (16.7) | 4 (57.1) |  |  |
|  | Severe | 1 (8.3) | 3 (42.9) |  |  |
| **P<sub>5</sub>** | Mild | 2 (66.7) | 0 (0.0) | 5.660† | **0.045** |
|  | Moderate | 1 (33.3) | 8 (88.9) |  |  |
|  | Severe | 0 (0.0) | 1 (11.1) |  |  |
| **p>5** | Mild | 5 (83.3) | 0 (0.0) | 6.407† | **0.024** |
|  | Moderate | 1 (16.7) | 3 (75.0) |  |  |
|  | Severe | 0 (0.0) | 1 (25.0) |  |  |

† Fisher's Exact test used.

Majority of the participants in both groups (90.9% vs 90.9%: p>0.999) were willing to come for repeat cervical smears in the future after undergoing initial cervical cancer screening. Although more participants in Gel group (98.5% vs 92.4%; p=0.208) were more satisfied with smear collection procedure than in the no Gel group, however, the difference was not statistically significant. This is shown in table 7.

**Table 7:** Satisfaction for Pap smear procedure and willingness for repeat of Pap smear among the study participants.

| Variables | | Study Group | | Total (%) | $\chi^2$ | p-value |
| --- | --- | --- | --- | --- | --- | --- |
|  |  | GEL (%) | No Gel (%) |  |  |  |
| **Satisfied with the PAP smear procedure?** | YES | 65 (98.5) | 61 (92.4) | 126 (95.5) | 2.794 | 0.208† |
|  | NO | 1 (1.5) | 5 (7.6) | 6 (4.5) |  |  |
|  | **Total** | 66 (100) | 66 (100) | 132 (100) |  |  |
| **Are you willing to come for repeat testing in future?** | YES | 60 (90.9) | 60 (90.9) | 120 (90.9) | 0.000 | >0.999 |
|  | NO | 6 (9.1) | 6 (9.1) | 12 (9.1) |  |  |
|  | **Total** | 66 (100) | 66 (100) | 132 (100) |  |  |

† Fisher's Exact test used.

Some participants were unwilling to repeat Pap smear test because the procedure was discomforting (33.3% vs 16.7%; p=0.691), painful (66.7% vs 66.7%; p=0.691) and very uncomfortable (0.0% vs 16.7%; p=0.691) in the Gel and No Gel group respectively. This is shown in table 8.

**Table 8:** Participants’ Reasons for unwillingness to repeat Pap smear test.

|  | **Gel (%)** | **No Gel (%)** | **Total (%)** | **Test value** | **p-value** |
| --- | --- | --- | --- | --- | --- |
| **Discomforting** | 2 (33.3) | 1 (16.7) | 3 (25.0) | 1.409 | >0.999† |
| **Painful** | 4 (66.7) | 4 (66.7) | 8 (66.7) |  |  |
| **Very uncomfortable** | 0 (0.0) | 1 (16.7) | 1 (8.3) |  |  |
| **Total** | 6 (100.0) | 6 (100.0) | 12 (100.0) |  |  |

† Fisher's Exact test used.

## Discussion

The practice of speculum lubrication for vaginal examination is varied due to the fears that lubricants can alter the adequacy of cytological smears. However, recent studies documenting the relationship between speculum lubrication and Pap smear accuracy have showed evidence supporting that speculum lubrication does not disrupt the evaluation of cytological smears.

Our findings revealed that there was no significant difference in the proportion of unsatisfactory cytology smear results in the gel group and no gel group (13.6% vs 21.2%; p=0.359) respectively. This was similar to the findings in previous studies.^10,11,18,19^ However; this was in contradiction to the findings of Charoenkwan et al and Kosus et al which showed statistically significant greater numbers of unsatisfactory smears in the participants that received speculum lubrication^20,21^. The reason for the differing report by Charoenkwan et al may be because; there was direct contamination of the smears with gel in their study as against speculum lubrication employed in our present study. Similarly, in Kosus et al, the risk of significantly higher inadequate results in the gel group could be because of the personnel that collected the smears as majority were inexperienced trainees.^21^ The experience of the provider has been implicated as one of the risk factors for smear inadequacy. ^11,18,19^, Also there are some reports in the literature that suggest that lubricants can affect the result of cervical cytology^22,23^ In most of these studies, lubricant gel was added intentionally to Liquid based cytology specimens. Most of these study designs do not reflect actual clinical practice.

The Bethesda system requires that Papanicolaou preparations must include enough cells to cover 10% of a slide.^18^ If 75% of the epithelial cells are obscured by blood, inflammation, or artifact, the slide is considered unsatisfactory.^18,19^ The reasons for the unsatisfactory smears in this study were scanty cells (87%) and obscurity by inflammation (13%). This finding was similar between the two groups. Gel overlay or obscurity by gel was not recorded as a reason for unsatisfactory results. This is similar to the findings by Amies et al and Gilson et al where the reason for unsatisfactory result was obscurity by blood, inflammation and gel overlay was not documented as a reason for unsatisfaction^10,18^.

This study has revealed that higher pain scores were recorded in the no gel group compared to the gel group (p<0.001). This finding agrees with previous studies by Uygur et al and Gungorduk et al where the pain scores were significantly higher in the arm that did not use speculum lubrication^11,24^. However, our findings differ from the work of Gilson et al that reported that speculum lubrication did not affect pain and discomfort^10^. The difference between the present study and that of Gilson et al may be because of the method of assessment of the pain used. In the study by Gilson et al, pain was assessed using Wong-Baker Faces

Pain Rating Scale which is less effective method of pain measurement. while in this study Numeric rating scale for pain which has been shown to be widely appropriate for pain evaluation globally was utilized.

All the samples came out negative for intraepithelial lesion or malignancy except one that was low grade squamous intraepithelial lesion. There was no diagnosis of invasive cancer or high grade squamous intraepithelial lesion. This absence or insignificant number of positive cytological reports of epithelial lesions has also been reported by other studies.^19,22^ This might be explained by the fact that study population was more of the asymptomatic younger age group, healthy volunteers and not the age group with higher risk of positive cytological reports of epithelial lesions.

When the pain scores assessed were compared according to parity of the participants, it was observed that the severity of the pains was seen at the extremes of parity. The reason for this peculiar finding may be because at nulliparous state the vagina and introitus may be narrower than in the parous state. However, the pain may also be more severe at higher parity because women with higher parity may have completed family size and are older or may be postmenopausal resulting in the vaginal atrophy and vaginitis. These conditions expectedly will elicit more pain reaction among the participants.

This study had revealed that more participants in Gel group (98.5% vs 92.4%; p=0.208) were more satisfied in the Pap smear collection procedure than in the no Gel group, but the difference was not statistically significant. In addition, equal proportion of the participants in each group (90.9% vs 90.9%; p>0.999) were willing to come for repeat cervical smears in the future after undergoing initial cervical cancer screening. This finding is not only interesting but encouraging. This could be explained by the fact that more than half (67.4%) of the study population had tertiary education, and 28% had secondary education resulting in a high level of awareness, knowledge and need for the uptake of the cervical screening among the study population. In a previous review by Chorley et al, it was revealed that a single negative experience prevented some women from re-attending screening, even if they had multiple positive previous experiences to draw upon. However, the studies involved in the Chorley et al systematic review were not randomized controlled trials and did not compare the failure at re-screening in women that received speculum lubrication versus no speculum lubrication.^25^

It was also noted that the reasons why some participants were unwilling to come for repeat testing in the future was due to pain (66.7%) and discomfort (25%). Although these findings do not differ among the two groups, it still helps to buttress the fact that pain while undergoing Pap smear collection is a strong reason why some women would not present for repeating testing.

The strength of the study was that it is a double blind randomized controlled study. The study also employed liquid based cytology thereby forming a literature base for future comparison as most of the studies in the literature employed the conventional Pap smear technique. However, the limitation was that only one type of water-based lubricant (KY-Jelly) was used, so results may not be applied to other kinds of lubricants. Also, our study only utilized metal speculums so the findings may also not be applied to plastic type of speculum. Perception of pain is complex and multifactorial, and so cultural, genetic and environmental factors may affect these results in different populations. The study was a single center based hence multi-center similar randomized studies are needed.

## Conclusion

In conclusion, speculum lubrication did not affect the adequacy of the cervical cytology smears, satisfaction for Pap smear procedure and the willingness to come for repeat cervical smears in the future after initial cervical cancer screening but significantly reduced the pain and discomfort experienced at Pap smear collection.

## Disclosure statement for publication

This work or its part has not been submitted to another journal for publications. All the authors contributed to conceptualization, or design, or drafting of manuscripts, or collection of data/data analysis and final drafting and revision of manuscript for submission.

## Funding

The researchers funded the research and none of the participants paid for the procedure or investigation

## Ethical considerations

The study adhered to CONSORT guidelines^26^. The approval for the study was obtained from the Ethics committee of Nnamdi Azikiwe University Teaching Hospital Nnewi (NAUTH), Nnewi, Nigeria on 24^th^ September 2019 with the approval reference number: NAUTH /CS/66/VOL.12/098/2019/040. The study was carried out in accordance with the ethical principles of clinical research involving human participants according to Helsinki declarations

## Consent to participate

All the recruited participants gave an informed written consent to participate in the study.

## Author contribution

Chito P. Ilika, the principal author, was involved in the conceptualization/design of the study, manuscript writing, revision, data collection. George U. Eleje; Michael E. Chiemeka, and Joseph I. Ikechebelu were involved in manuscript writing, revision and supervision. Chigozie G. Okafor; Emmanuel O. Ugwu, Ifeanyichukwu, J. Ofor, Onyecherelam M. Ogelle, Osita S. Umeononihu, Johnbosco E. Mamah, Chinedu L. Olisa, Chijioke O. Ezeigwe, Malarchy E. Nwankwo and Chukwuemeka J. Ofojebe participated in manuscript writing and revision. Frances N. Ilika; Valentine C. Ilika; Chidinma C. Okafor, Onyeka C. Ekwebene and Obinna K. Nnabuchi participated in manuscript writing, revision and Data analysis.

## Trial Registration

The Trial was registered with Pan-African-Clinical-Trial-Registry on 08/07/2020 with unique identification/registration-no: PACTR2020077533364675.

## Data Availability

Data is available upon request from the authors

## Acknowledgment

The authors sincerely appreciate everyone that contributed to the success of this study especially the patients that were recruited into the study and the NAUTH, Nnewi, Nigeria.

## Conflict of interest

There was no conflict of interests.

## Notes

### Competing Interest Statement

The authors have declared no competing interest.

### Funding Statement

The author(s) received no specific funding for this work.

### Author Declarations

The approval for the study was obtained from the Ethics committee of Nnamdi Azikiwe University Teaching Hospital Nnewi (NAUTH), Nnewi, Nigeria on 24th September 2019 with the approval reference number: NAUTH /CS/66/VOL.12/098/2019/040. The study was carried out in accordance with the ethical principles of clinical research involving human participants according to Helsinki declarations

